# Black/African American Communities are at Highest Risk of COVID-19: Spatial Modeling of New York City ZIP Code-Level Testing Results

**DOI:** 10.1101/2020.05.14.20101691

**Authors:** Charles DiMaggio, Michael Klein, Cherisse Berry, Spiros Frangos

## Abstract

**Introduction:** The population and spatial characteristics of COVID-19 infections are poorly understood, but there is increasing evidence that in addition to individual clinical factors, demographic, socioeconomic and racial characteristics play an important role.

**Methods:** We analyzed positive COVID-19 testing results counts within New York City ZIP Code Tabulation Areas (ZCTA) with Bayesian hierarchical Poisson spatial models using integrated nested Laplace approximations.

**Results:** Spatial clustering accounted for approximately 32% of the variation in the data. For every one unit increase in a scaled standardized measure of Chronic Obstructive Pulmonary Disease (COPD) in a community, there was an approximate 8-fold increase in the risk of a positive COVID-19 test in a ZCTA (Incidence Density Ratio = 8.2, 95% Credible Interval 3.7, 18.3). There was a nearly five-fold increase in the risk of a positive COVID-19 test. (IDR = 4.8, 95% Cr I 2.4, 9.7) associated with the proportion of Black / African American residents. Increases in the proportion of residents older than 65, housing density and the proportion of residents with heart disease were each associated with an approximate doubling of risk. In a multivariable model including estimates for age, COPD, heart disease, housing density and Black/African American race, the only variables that remained associated with positive COVID-19 testing with a probability greater than chance were the proportion of Black/African American residents and proportion of older persons.

**Conclusions:** Areas with large proportions of Black/African American residents are at markedly higher risk that is not fully explained by characteristics of the environment and pre-existing conditions in the population.

## Introduction

The SARS-Cov-2 virus poses unprecedented clinical and public health challenges world-wide. While much of the attention has been rightfully focused on the clinical aspects of the disease, epidemiological studies and prevention research are becoming of increasing importance, particularly as no effective therapeutic has yet been identified.^1^ Epidemiological and population-based studies can contribute to the identification of patient risk factors for disease severity. Recent studies of observational registry data have found COVID-19 mortality to be independently associated with coronary artery disease (CAD) (Odds Ratio (OR) for mortality=2.7, 95% CI 2.1, 3.5), chronic obstructive pulmonary disease (COPD) (OR =3.0; 95% CI, 2.0 to 4.4), and age greater than 65 years (OR = 1.9; 95% CI, 1.6 to 2.4).^2^ In one case series, 68% of laboratory-confirmed COVID-19 ICU patients had at least one comorbidity, of which hypertension was most common.^3^

Not all risks, however, are physiologic. As the COVID-19 pandemic continues to ravage communities across the United States and the world, attention is increasingly turning to population-level demographic, socioeconomic, racial and environmental risk factors for COVID- 19. Blacks/African Americans have been reported to contract and die from COVID-19 at higher rates than others.^4^ In Chicago, a large number of COVID-19 deaths are concentrated in five largely black neighborhoods.^5^ A similar mortality concentration among Black/African American persons has been reported in New Orleans.^6^ At the built-environmental level, drivers of disease include population density ^7^ and housing density, with urban counties in the US having the highest COVID-19 death rates.^8^

Few regions of the US have been more grievously affected than the five boroughs of New York City. A neighborhood-level analysis of New York City found higher rates of COVID-19 disease in areas with higher population shares of Black/African American and Hispanic persons, and in areas with higher population density.^9^ While it certainly is possible that those affected have higher rates of underlying health conditions that may increase their susceptibility to the virus, the authors speculate that “residents of these neighborhoods are less likely to be able to work from home, disproportionately rely on public transit during the crisis, are less likely to have internet access,” and “have higher rates of overcrowding at the household level.”

In this analysis, we analyze positive COVID-19 testing result counts within New York City ZIP Code Tabulation Areas (ZCTA) using Bayesian hierarchical Poisson spatial models with integrated nested Laplace approximations. We attempt to quantify the amount of spatial clustering in New York City neighborhoods, and the association of positive test counts in a neighborhood with population-level estimates of demographic, socioeconomic, health, and built environmental variables. The results quantify and provide insights into the complex interplay of individual and ecologic risks for COVID-19 spread and may be helpful in the effective allocation of testing resources and interventions in similar urban settings.

## Methods

### Data

COVID-19 test result data were obtained from the New York City Department of Health and Mental Hygiene (NYC DOHMH) GitHub Page. Variables consisted of ZIP Code Tabulation Area (ZCTA) designation, total number of positive test and total number of tests performed. Files are updated approximately every 2 days. The data in these analyses were current as of 22 April 2020.

ZCTA-level data for total population, proportion of persons older than 65, number of persons self-identifying as Black/African American, Asian or Hispanic, number of persons older than 5 speaking a language other than English, population density, housing density, school density, (number of people, housing structures and schools per square mile respectively), proportion of persons receiving public assistance, were obtained from or derived from the US Census.^10^

We created a social fragmentation index based on the work of Congdon^11^ which combines 4 variables extracted from US census variables: the proportion of total housing units in a ZCTA that are not owner occupied, the proportion of vacant housing units, the proportion of individuals living alone, and the proportion of units into which someone recently moved. Based on Census definitions, a “recent” move is defined as anytime in the previous 9 years (since the last decennial census). Variables are standardized and added with equal weight. The resulting variable is normally distributed with mean zero and 95% quantiles -2.463311 and 2.205669.

Data on ZCTA health metrics were derived from shapefiles downloaded from the Simply Analytics company ^12^ and consisted of the number of persons in a ZCTA with heart disease or congestive heart failure (which are combined as a metric) and the number of persons with COPD.

Spatial shapefiles of New York City ZCTAs were downloaded and derived from the New York City Department of City Planning.^13^ The testing and covariate data were merged to the spatial shapefile data and restricted to ZCTAs with valid data entries. An adjacency matrix was created from the map file using the R tool `spdep::poly2nb()`, and manually edited to create adjacencies between New York City boroughs using `spdep::edit.nb()`.

### Statistical Analysis

After merging the testing to the covariate data, descriptive statistics consisted of counts, means and medians and maps of the number of positive COVID-19 tests per 10,000 total population and 10,000 tests performed in a ZCTA.

Counts of positive COVID-19 test results in New York City ZCTAs were spatially modeled according to Besag-York-Mollie as described by Lawson.^14–16^

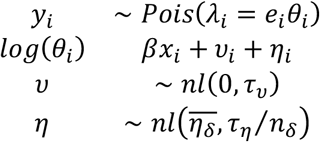

where,

1. the *y_i_* counts in area *i*, are independently identically Poisson distributed and have an expectation in area *i*. of *e_i_*, the expected count, times *θ_i_*, the risk for area *i*.
2. a logarithmic transformation (*log(λ_i_*)) allows a linear, additive model of regression terms (*βx_i_*), along with
3. a spatially random effects component (υ*_i_*) that is i.i.d normally distributed with mean zero (~ *nl*(0*,τ_η_))*, and
4. a conditional autoregressive spatially structured component 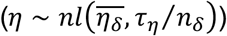 in which a “neighborhood” consisting of spatially adjacent shapes is characterized by the normally distributed mean of the spatially structured random effect terms for the spatial shapes that make up the neighborhood 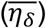, and the standard deviation of that mean divided by the number of spatial shapes in the neighborhood (*τ_η_/n_δ_*). This spatially structured conditional autoregression component is also sometimes described as a Gaussian process *λ* ~ (W, *τ_λ_*) where W represents the matrix of neighbors that defines the neighborhood structure, and the conditional distribution of each *λ_i_*, given all the other *λ_i_* is normal with *μ* = the average *λ* of it,s neighbors and a precision(*τ_λ_*).

A baseline convolution model that consisted solely of an intercept term with unstructured and spatially structured random effect terms was extended to include univariate association of explanatory variables with the number of positive COVID-19 tests in a ZCTA. Important and likely associations were chosen for inclusion in a multivariable model with the primary exposure variable being the proportion of Black/African American residents in an area and additional explanatory variables included as potential confounders.

The final linear model consisted of an intercept (*β*_0_); a vector of scaled ZCTA-level explanatory variables 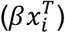 for the proportion of persons in a ZCTA identifying as Black/African American, with COPD, heart disease, older than 65 years, a measure of housing density, a spatially unstructured random effect term (*υ_i_*), and a spatially structured conditional autoregression term (*η_i_*). An offset varible for the total number of tests was included in all models. Model selection was based on deviance information criteria and number of effective parameters.

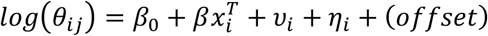

The spatially unstructured random effect term captures normally-distributed or Gaussian random variation around the mean or intercept. The spatially-structured conditional autoregression term accounts for local geographic influence. The intercept is interpreted as the average city-wide risk on the log scale adjusted for the covariates, random effects and spatial terms. The exponentiated coefficients for the explanatory covariates are interpreted as incidence density ratios. Coefficient results are presented with 95% Bayesian Credible Intervals (95% Cr I)

Spatial risk, controlling for or holding the covariates constant, was calculated as 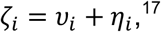 and is interpreted as the residual spatial risk for each area (compared to all of New York City) after covariates and spatial clustering are taken into account. The probability of spatial risk greater than 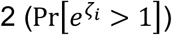 was calculated. As originally described by Clayton and Bernardinelli,^18^ these exceedance probabilities are the posterior probabilities for an area’s spatial risk estimate exceeding some pre-set value. This was extended by Richardson, et al ^19^ to decision rules “for classifying whether (an area) has an increased risk based on how much of the posterior distribution of the relative risk parameter … exceeds a reference threshold”.^20^ They are calculated as the proportion of simulations for which the linear combination of effects 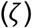 exceeds the target value. Lastly, the proportion of spatially explained variance was calculated as the proportion of total spatial heterogeneity accounted for by the spatially structured conditional autoregression variance.^17^

Spatial modeling was conducted using integrated nested Laplace approximations (INLA) with the R INLA package^21^ using approaches described by Blangiardo, et al. ^17^ The study protocol was exempted as not human research by the New York University School of Medicine Institutional Review Board.

## Results

### Descriptive Statistics

There were 177 ZCTA’s in the data set. The mean COVID-19 positive test rate per 10,000 ZCTA population was 166.2 (95% CI 156.7, 175.7). The mean COVID-19 positive test rate per 10,000 tests was 5,176.0 (95% CI 5,045.9, 5,306.1) and appeared skewed and peaked, indicating that a relatively small number of ZCTAs accounted for highest rates. (Figure 1) The 5 ZCTAs with highest positive COVID-19 test numbers per 10,000 population were the same as those with the highest proportion per 10,000 tests (10464, 10470, 10455, 10473, 11234, and 11210). The 5 lowest ZCTAs were also the same for both measures (11103, 11102, 11693, 11369, 11363, and 10308). Table 1 presents comparative statistics for the ZCTA’s with the highest and lowest quantiles for population-based positive test rates. Figure 2 presents a choropleth of positive COVID-19 tests per 10,000 per 10,000 positive tests.

**Figure 1.**
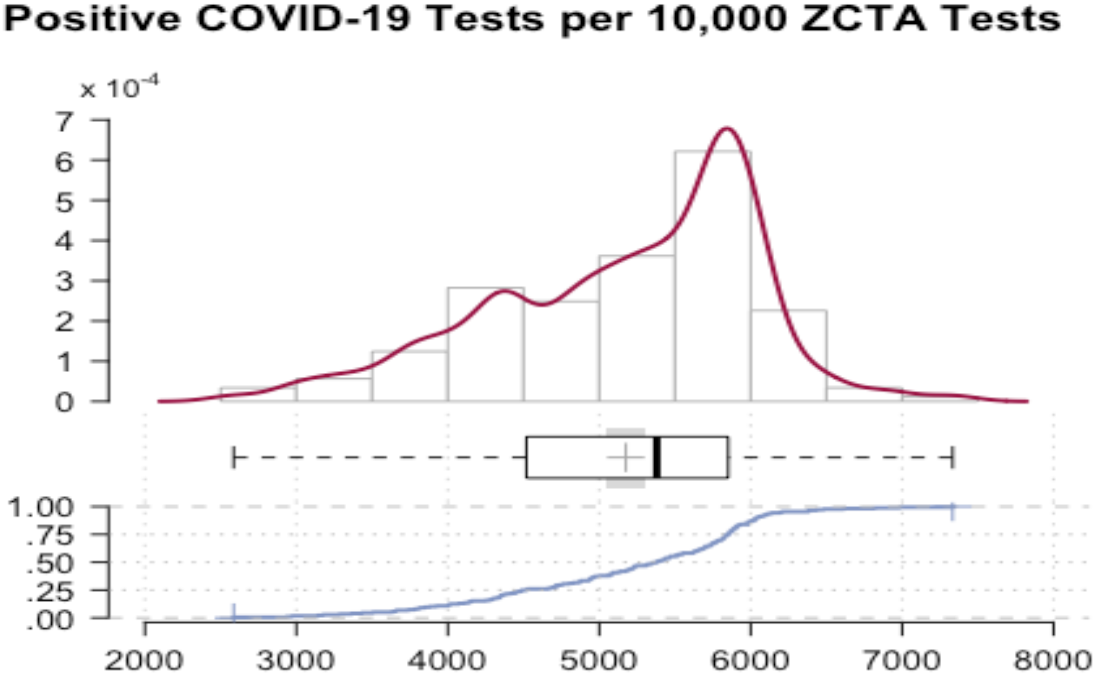
COVID-19 Positive Test rate per 10,000 tests. New York City, April 3- 22, 2020.

**Figure 2.**
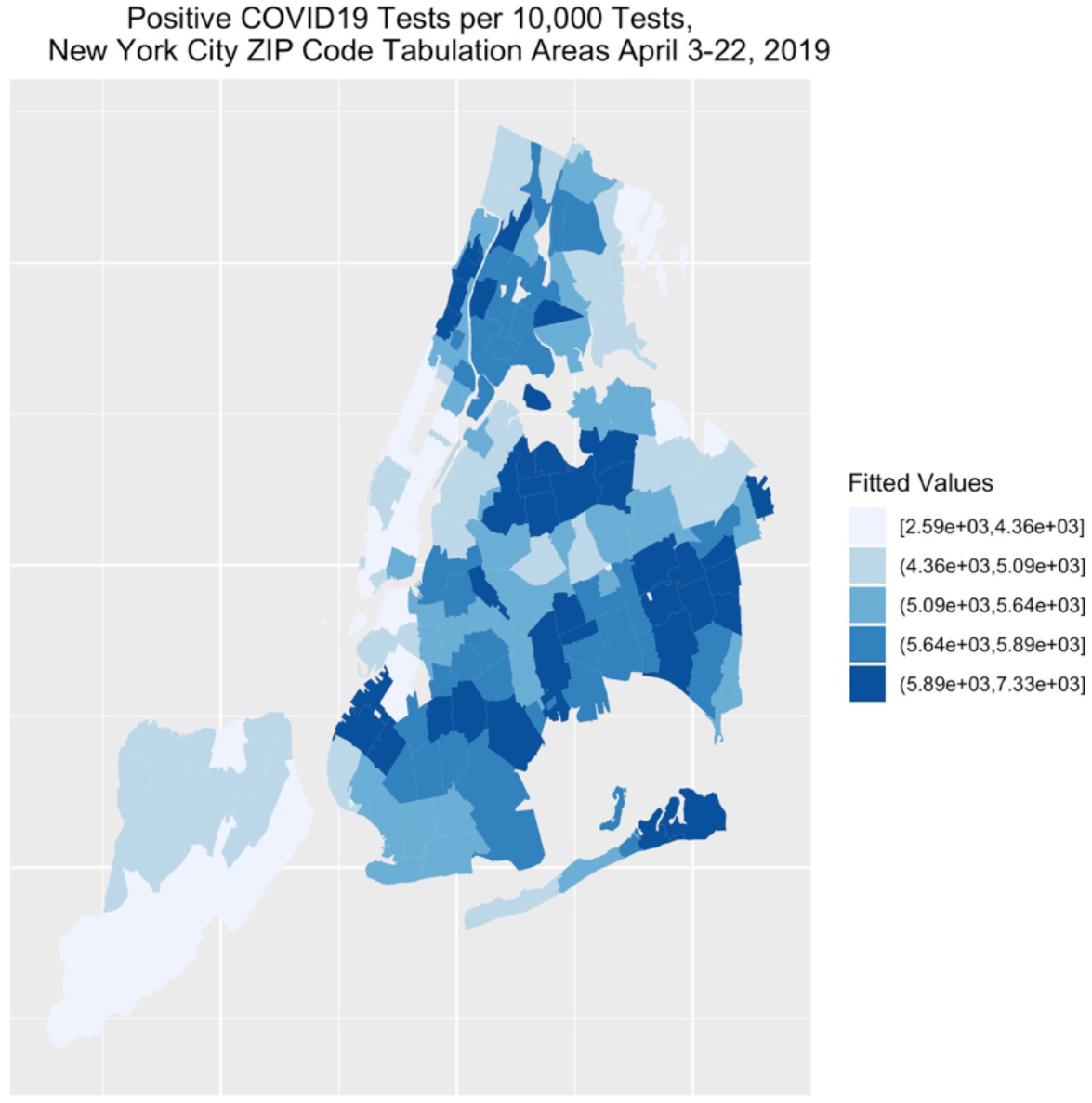
Choropleth Quintiles Positive COVID-19 Tests per 10,000 Tests. New York City, April 3- 22, 2020.

**Table 1.**
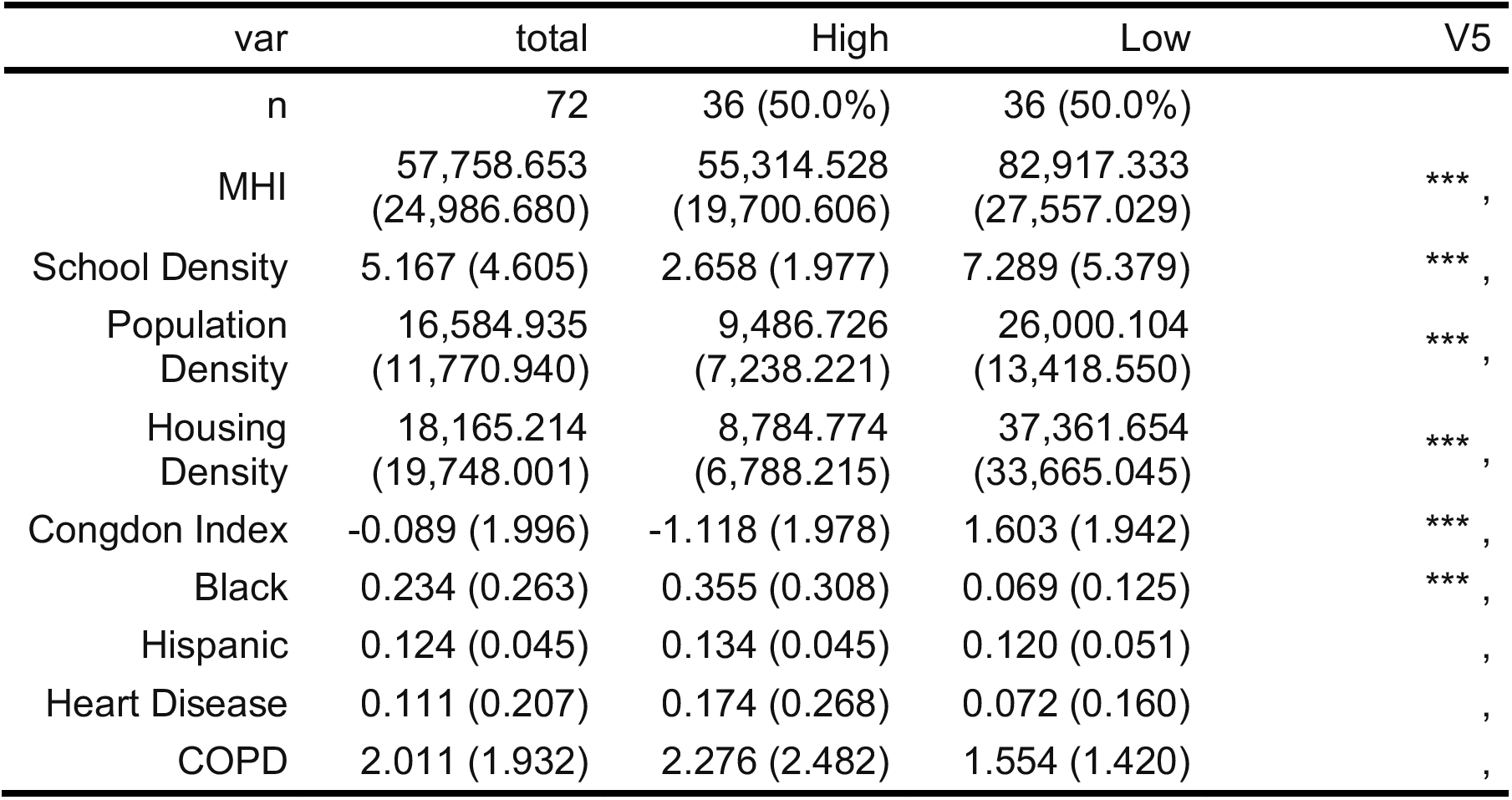
Comparative Statistics Positive COVID-19 Tests per 10,000 Population, High and Low Quantile ZIP Code Tabulation Area. New York City, April 3- 22, 2020.

### Spatial Models

A frailty model consisting of only a random effect term and no explicit spatial component, returned a deviance information criteria (DIC) 1831.58, with 174.5 effective parameters. The random effect term was normally distributed around the mean value of 64.9 (sd= 1.1, 95% Credible Interval 55.5, 75.6) reflecting random nature of the distribution of the unstructured heterogeneity or variance.

A convolution model with a spatially-structured conditional autoregression term added to the spatially-unstructured heterogeneity random effect term of the frailty model, returned a DIC of 1807.60 (with 175.98 effective parameters) reflecting an improvement over the baseline unstructured heterogeneity frailty model, and indicating the spatial component added information to the simple unstructured model. In Figure 3, the spatial risk estimate is calculated as the sum of the unstructured and spatially structured variance components 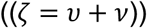 Finally, in figure 4, we calculate and map the probability of relative risk greater than 1, which is interpreted in the context of figure 3 as a type of “hot spot” map place the risk estimates in the context of their probabilities. Lastly, we estimate the proportion of the variance explained by geographic variation or place, which for this model is approximately 32%

**Figure 3.**
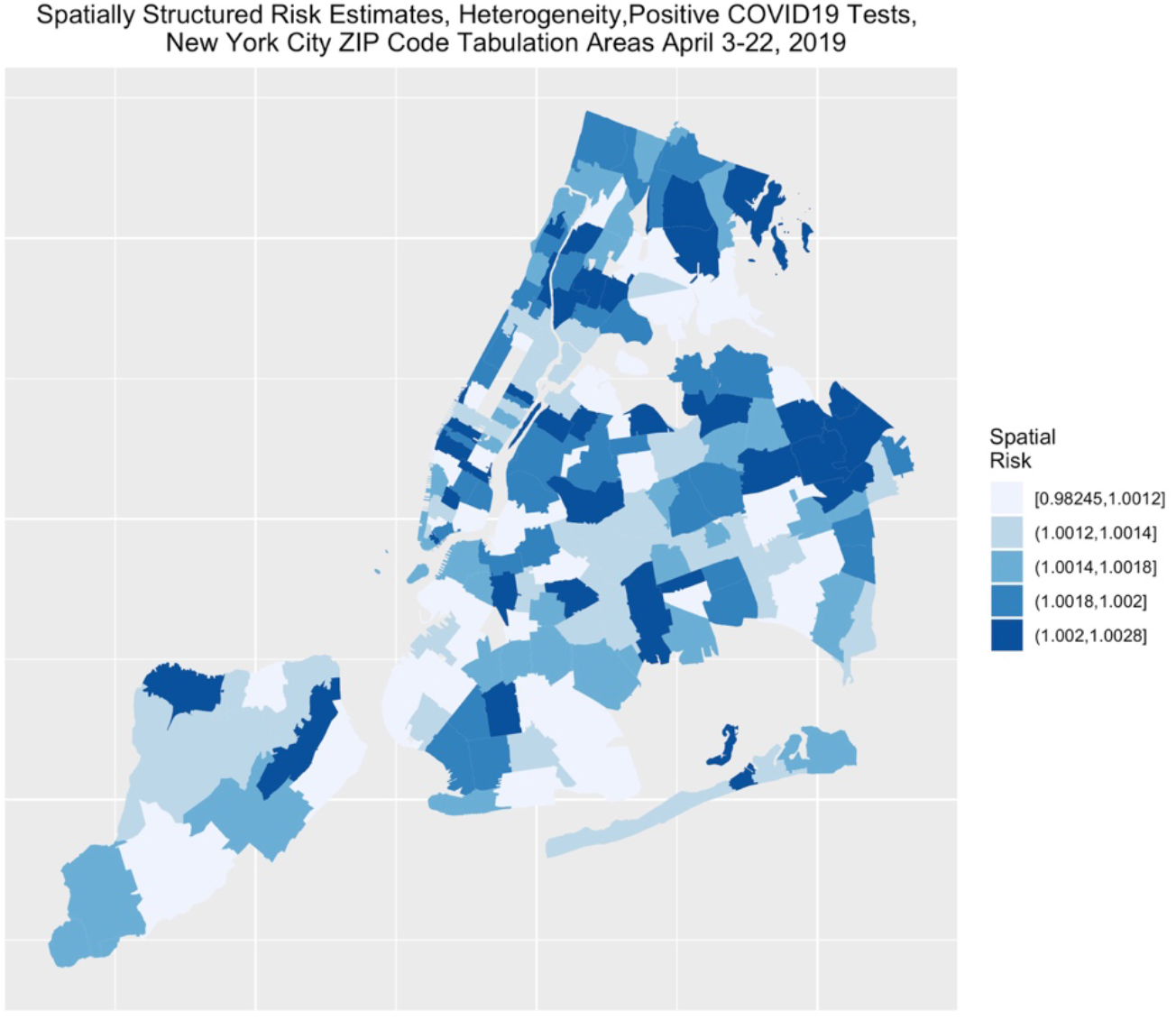
Choropleth Quintiles Spatial Risk Estimates (Sum of Unstructured and Spatially Structured Variance) Positive COVID-19 Tests per 10,000 Tests. New York City, April 3- 22, 2020.

**Figure 4.**
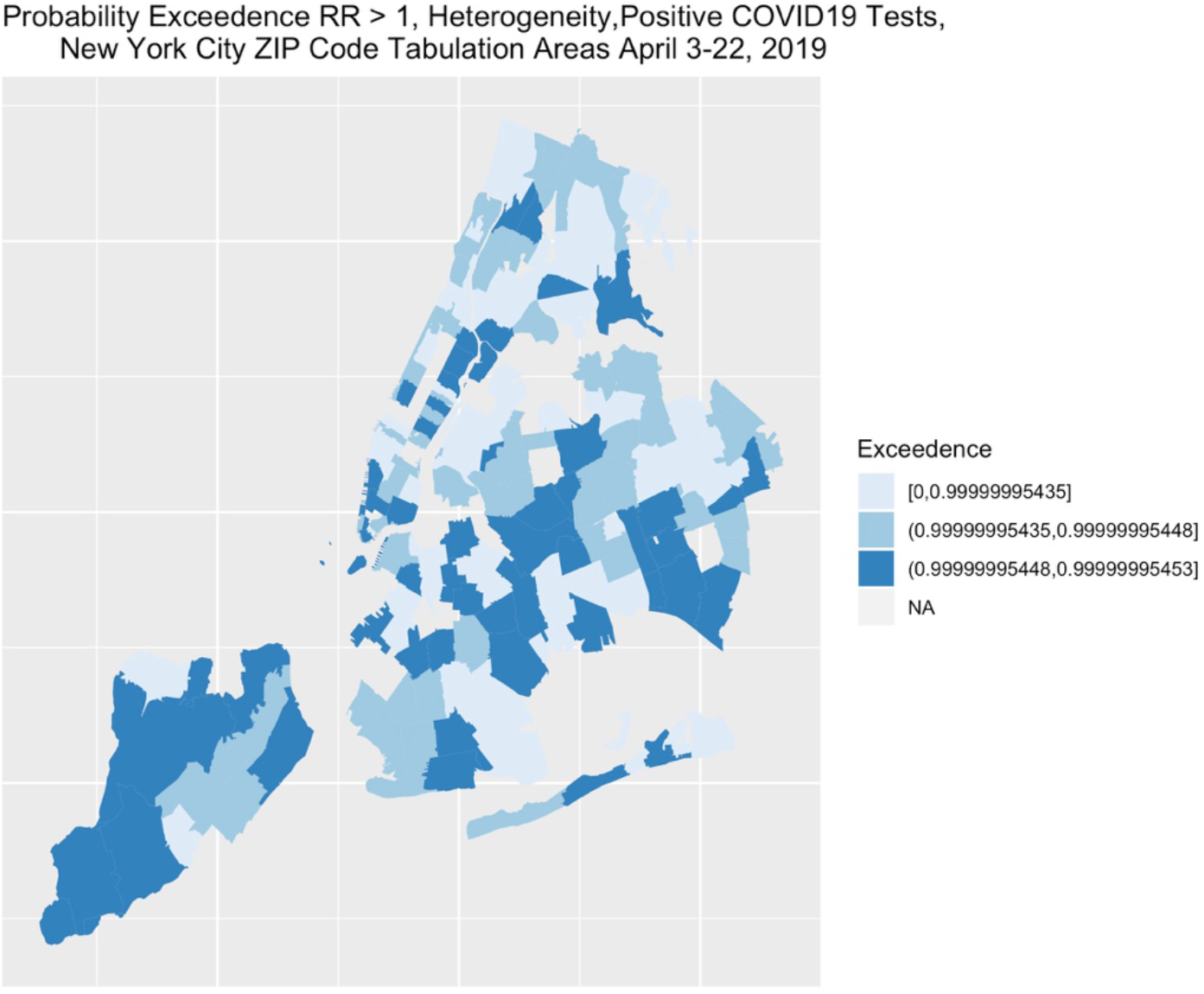
Choropleth Three Equal Groups, Probability of Relative Risk Greater than 1. Positive COVID-19 Tests per 10,000 Tests. New York City, April 3- 22, 2020.

### Simple and Multivariable Models

The convolution model is extended to include ecological-level variables for population, housing, income, social fragmentation, population characteristics, and clinical conditions. Table 2 summarizes the results of a series of unadjusted single Covariate Models of Associations with Positive COVID-19 Test Counts. The single strongest unadjusted association is for the proportion of persons in a ZCTA with COPD, which returned an incidence density ratio (IDR) of 8.2 (95% Credible Interval 3.7, 18.3), indicating that for each single unit increase in the standardized proportion of persons in a ZCTA with COPD, there was an eight-fold increased risk of an additional positive COVID-19 test in that ZCTA. The proportion of Black/African American residents in a ZCTA was also strongly associated with the risk of positive COVID-19 tests. For every one unit increase in a scaled standardized measure of the proportion of Black/African American residents, there was a nearly five-fold increase in the risk of a positive COVID19 test. (IDR = 4.8, 95% Cr I 2.4, 9.7)

**Table 2.**
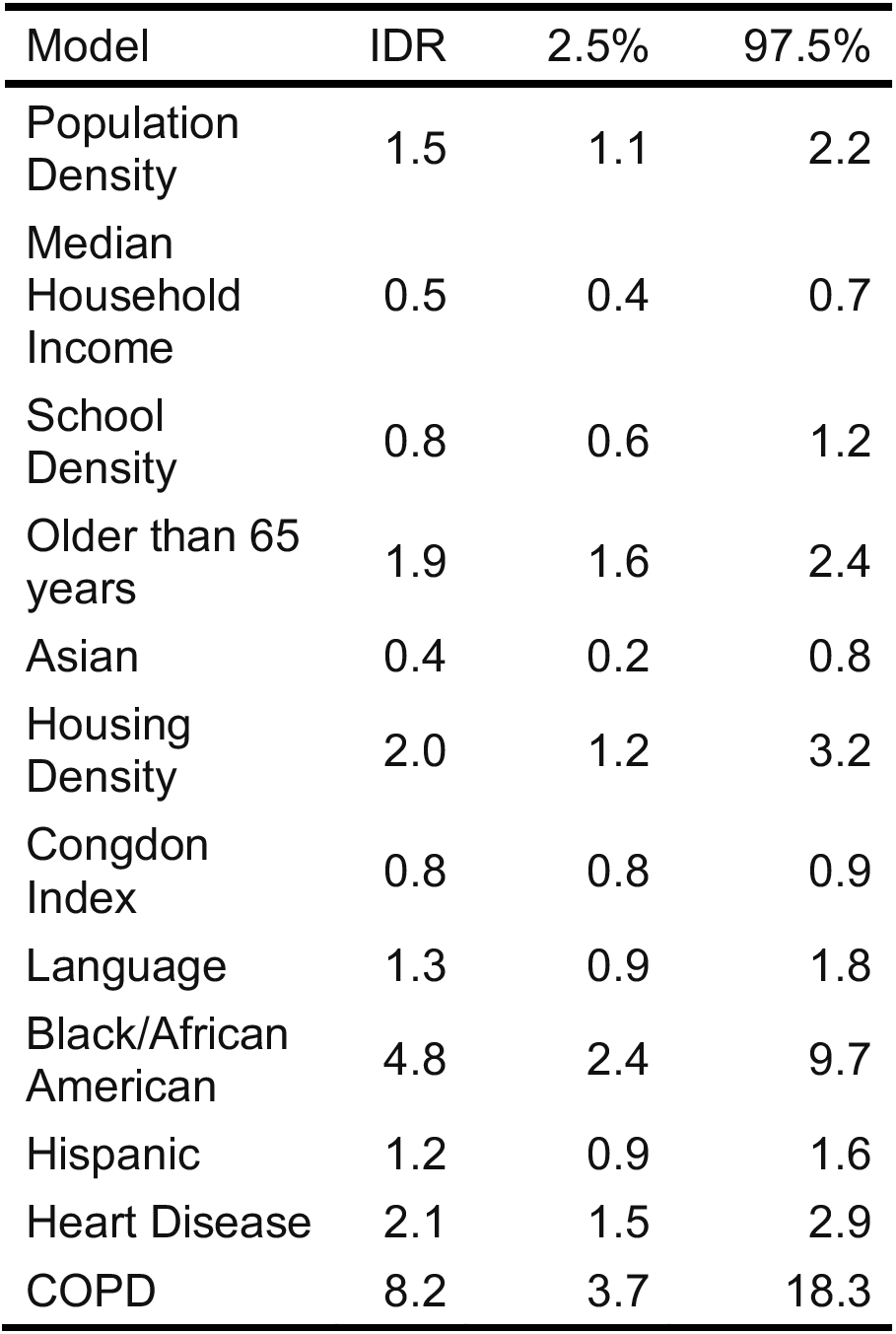
Summary Series of Unadjusted Single Covariate Bayesian Hierarchical Poisson Models for Association with Positive COVID-19 Tests Counts in New York City ZIP Code Tabulation Areas, April 3- 22, 2020.

**Table 3.** Summary Multivariable Bayesian Hierarchical Poisson Modes for Association with Positive COVID-19 Tests Counts in New York City ZIP Code Tabulation Areas, April 3- 22, 2020.

| Variable | IDR | 2.5% | 97.5% |
| --- | --- | --- | --- |
| Intercept | 353.82 | 197.66 | 632.23 |
| COPD | 2.32 | 0.92 | 5.85 |
| Heart Disease | 1.27 | 0.88 | 1.83 |
| Black/African American | 2.29 | 1.13 | 4.68 |
| Older than 65 years | 1.50 | 1.17 | 1.92 |
| Housing Density | 1.08 | 0.65 | 1.78 |

Variables for population density, proportion of residents older than 65 years, housing density, and heart disease were also associated with increased risk of positive COVID-19 testing rates. Median household income (MHI) in a ZCTA community was inversely related to positive COVID- 19 tests. For each unit increase in a standardized measure of MHI in a ZIP Code Tabulation Area, there is an approximately 46% *decrease* in the number of positive COVID19 tests. (Incidence Density Ratio = 0.54, 95% CrI 0.43, 0.69). Other variables that were associated with lower positive tests were proportion of Asian and proportion of Hispanic residents and increased measures of social fragmentation. School density, proportion of persons not speaking English, and the proportion of persons on public assistance were not associated with positive COVID-19 testing rates.

In a multivariable model including COPD, heart disease, proportion of Black/African American residents, housing density, and age greater than 65 years, the only 2 variables that remained associated with positive COVID-19 testing with a probability greater than chance were the proportion of Black/African American residents and older persons. (Table 4) Proportion of Black/African American residents was the strongest predictor of higher positive testing rates in a community regardless of other factors.

## Discussion

Despite the recent onset of the current COVID-19 pandemic, there is already growing evidence about both individual risk factors and population-level drivers of disease and mortality. This study adds to the number of very recent similar spatial analyses of ZCTA-level testing data released by the New York City Department of Health and Mental Hygeiene,^22-24^ and illustrates the importance of sharing these kinds of data, as well as the informative nature of spatial epidemiology as the pandemic evolves across the nation and the world. Consistent with prior reports, we find that the clustering of positive COVID-19 testing results in NYC are unlikely to be due to chance,^9, 23^ and is driven in large measure by socioeconomics, age distribution,^24^ and race.^9, 23^

Our study adds to this by demonstrating that the proportion of residents self-identifying as Black/African American is among the single strongest unadjusted bivariate predictors of the proportion of positive tests in a community. The only stronger such predictor is the proportion of residents with COPD, which at 8 times the risk of areas with less COPD, is stunning. But perhaps the more unexpected finding is that when Black/African American race and COPD are considered jointly, it is race that appears to be the stronger predictor. Unlike a previous New York City-based report,^9^ we did not find an independent risk associated with the proportion of Hispanic residents. It may be that census estimates of Black/African American persons includes persons who also identify as Hispanic. Three of the 5 ZCTAs with highest positive COVID-19 test numbers per 10,000 population were in areas of the Bronx with large proportions of Hispanic and Latino residents. And, it may be that disparities may vary depending in part on how well-established Hispanic communities are within cities and states. ^25^

The question of why COVID-19 affects one community more severely than another may provide clues to crucial questions about who is a risk and why.^26^ Our study indicates place is important. We find about a third of the variance in a simple spatial model can be accounted for by place. We found risk to be approximately doubled by environmental characteristics like population and housing density. This complements a report of a non-spatial, linear multivariable regression model of similar data that reported that 72% of variance could be attributed to individual characteristics like household size, gender, age, race and immigration status.^22^.

If ecologic and spatial analyses can provide clues, it remains to be determined what those clues point to. It could be that Black/African American race is a proxy for underlying physiological risks. There are preliminary reports that infection with SARS-Cov-2 may be associated with the A blood group,^27^ and that severe COVID-19 is driven in part by coagulopathies that may be associated with Factor VIII and von Willebrand factor,^28^ although the relationship of such factors with race is complex.^29^ Race may also be associated with exposure. By one account, 80% of non-medical staff in New York City’s hard-hit public hospitals are Black/African American or Hispanic, compared to less than half of doctors and nurses. It may be that their risk of exposure to SARS-Cov-2 and need for similar personal protective equipment has been underestimated.^30^ Our results indicate that such exposure may be a more critical factor than underlying health conditions. That a risk like COPD should drop from a level of 8 to nonsignificance when race is included in the model may point to something beyond physiological risk of infection. Placing risk factors in context, both within and across populations, may be key. Differences between New York City vs Chicago as a whole appear to influence the relative strength of associations between population-level demographic variables and testing outcomes.^23^ It will be increasingly important to conduct such comparative studies.

Our results should be interpreted cautiously. Ecological studies can offer a view of disease processes in a community, but it may be a fractured view. Measures like school density and social fragmentation may not be measuring what we think they are measuring; the number of schools in an area, rather than acting as a disease multiplier, may be a measure of the strength of the tax base. Similarly, the Congdon Index treats empty houses as a measure of disorder, but this has a very different meaning in the setting of a rapidly-spreading infectious disease. The proportion of non-English speakers in a given ZCTA may be biased by a lack of self-reporting by undocumented immigrants. And, as in any ecologic study, it is not certain that the persons with the risk factor being studied are those who are developing the outcome.

SARS-Cov-2 testing results are imperfect, with numbers likely to be biased by the availability of testing. But, we would expect that bias, to be in the direction of increased counts in areas with higher socioeconomic status. Consistent with our findings, a recent geographic analysis reported that persons in poorer NYC neighborhoods were less likely to be tested but once tested were more likely to test positive.^22^ It is partly for this reason, we chose to base most of our analyses on the proportion of positive tests, rather than the population-based rates of positive tests, an approach taken by others. ^22^

Despite these caveats, it is difficult to overlook the interplay of race and COVID-19. Race appears to be an indicator of risk independent of social status, income, built environment or even underlying health. This has implications not only for justice and equity, but for an effective response to the pandemic.

## Data Availability

Data is publicly available.

## References

1. Information for Clinicians on Investigational Therapeutics for Patients with COVID-19. Centers for Disease Control and Prevention: Coronavirus Disease 2019 (COVID-19). Published online April 25, 2020. https://www.cdc.gov/coronavirus/2019-ncov/hcp/therapeutic-options.html

2. Mehra MR, Desai SS, Kuy S, Henry TD, Patel AN. Cardiovascular disease, drug therapy, and mortality in covid-19. N Engl J Med. Published online May 2020. doi:10.1056/NEJMoa2007621

3. Grasselli G, Zangrillo A, Zanella A, et al. Baseline characteristics and outcomes of 1591 patients infected with SARS-Cov-2 admitted to ICUs of the Lombardy region, Italy. JAMA. Published online April 2020. doi:10.1001/jama.2020.5394

4. Yancy CW. COVID-19 and African Americans. JAMA. Published online April 2020. doi:10.1001/jama.2020.6548

5. Reyes C GC Husain N. Chicago,s coronavirus disparity: Black Chicagoans are dying at nearly six times the rate of white residents, data show. Chicago Tribune. Published online 2020. https://www.chicagotribune.com/coronavirus/ct-coronavirus-chicago-coronavirus-deaths-demographics-lightfoot-20200406-77nlylhiavgjzb2wa4ckivh7mu-story.html. Accessed 9 May 2020

6. M D. Louisiana data: Virus hits Blacks, people with hypertension. US News World Report. Published online 2020. https://www.usnews.com/news/best-states/louisiana/articles/2020-04-07/louisiana-data-virus-hits-blacks-people-with-hypertension Accessed 9 May 2020

7. CDC COVID-19 Response Team. Geographic differences in covid-19 cases, deaths, and incidence - united states, February 12 - April 7, 2020. MMWR Morb Mortal Wkly Rep. 2020;69(15):465–471. doi:10.15585/mmwr.mm6915e4

8. R F. The geography of coronavirus. Bloomberg CityLab. Published online 2020. https://www.citylab.com/equity/2020/04/coronavirus-spread-map-city-urban-density-suburbs-rural-data/609394/ Accessed 9 May 2020

9. COVID-19 cases in New York City, a neighborhood-level analysis. The Stoop: NYU Furman Center Blog. Published online 2020. https://furmancenter.org/thestoop/entry/covid-19-cases-in-new-york-city-a-neighborhood-level-analysis Accessed 9 May 2020

10. US Census Bureau. Washington, DC: 2010. https://data.census.gov/cedsci/ Accessed 9 May 2020

11. Congdon P. Bayesian models for suicide monitoring. European J Population. 2001;15(3):1–34.

12. SimplyAnalytics. https://simplyanalytics.com/ Accessed 9 May 2020

13. City Planning NYCD of. Bytes of the big apple. Published online 2011. https://www1.nyc.gov/site/planning/data-maps/open-data.page Accessed 9 May 2020.

14. Besag J, York J, Mollie A. Bayesian image restoration, with two applications in spatial statistics. Ann Inst Statist Math. 1991;43(1):1–59.

15. Lawson A, Biggeri A, Boehning D, et al. Disease mapping models: An empirical evaluation. Disease mapping collaborative group. Stat Med. 2000;19(17-18):2217-2241.

16. Lawson AB. Bayesian Disease Mapping: Hierarchical Modeling in Spatial Epidemiology, Second Edition (Chapman & Hall/CRC Interdisciplinary Statistics). Chapman; Hall/CRC; 2013:396.

17. Blangiardo M, Cameletti M, Baio G, Rue H. Spatial and spatio-temporal models with r-INLA. Spat Spatiotemporal Epidemiol. 2013;7:39-55.

18. Clayton D, Bernardinelli L. Bayesian methods for mapping disease risk. Geographical and environmental epidemiology: methods for small area studies. Published online 1992:205-220.

19. Richardson S, Thomson A, Best N, Elliott P. Interpreting posterior relative risk estimates in disease-mapping studies. Environ Health Perspect. 2004;112(9):1016–1025.

20. Best N, Richardson S, Thomson A. A comparison of Bayesian spatial models for disease mapping. Stat Methods Med Res. 2005;14(1):35–59.

21. Rue H, Martino S, Lindgren F, Simpson D, Riebler A. INLA: Functions which allow to perform full bayesian analysis of latent gaussian models using integrated nested Laplace approximation. Published online 2013. https://rdrr.io/github/andrewzm/INLA/man/INLA-package.html Accessed 9 May 2020

22. Borjas GJ. Demographic Determinants of Testing Incidence and Covid-19 Infections in New York City Neighborhoods. National Bureau of Economic Research; 2020. https://www.hks.harvard.edu/publications/demographic-determinants-testing-incidence-and-covid-19-infections-new-york-city Accessed 9 May 2020

23. Maroko AR, Nash D, Pavilonis B. Covid-19 and inequity: A comparative spatial analysis of New York City and Chicago hot spots. medRxiv. https://www.medrxiv.org/content/10.1101/2020.04.21.20074468v1 Published online 2020. Accessed 9 May 2020

24. Whittle RS, Diaz-Artiles A. An ecological study of socioeconomic predictors in detection of covid-19 cases across neighborhoods in New York City. medRxiv. https://www.medrxiv.org/content/10.1101/2020.04.17.20069823v1 Published online 2020. Accessed 9 May 2020

25. Jordon M and Oppel RA. For Latinos and COVID-19, Doctors are Seeing an “Alarming” Disparity. New York Times. 7 May 2020 https://www.nytimes.com/2020/05/07/us/coronavirus-latinos-disparity.html?referringSource=articleShare. Accessed 9 May 2020

26. Beech H KA Rubin AJ, R M. The covid-19 riddle: Why does the virus wallop some places and spare others? New York Times. Published online May 2020. https://www.nytimes.com/2020/05/03/world/asia/coronavirus-spread-where-why.html Accessed 9 May 2020

27. Zhao J, Yang Y, Huang H-P, et al. Relationship between the ABO blood group and the covid-19 susceptibility. medRxiv. https://www.medrxiv.Org/content/10.1101/2020.03.11.20031096v2 Published online 2020. Accessed 9 May 2020

28. Panigada M, Bottino N, Tagliabue P, et al. Hypercoagulability of covid-19 patients in intensive care unit. A report of thromboelastography findings and other parameters of hemostasis. J Thromb Haemost. Published online April 2020. doi:10.1111/jth.14850

29. Miller CH, Dilley A, Richardson L, Hooper WC, Evatt BL. Population differences in von willebrand factor levels affect the diagnosis of von willebrand disease in african-american women. Am J Hematol. 2001;67(2):125–129. doi:10.1002/ajh.1090

30. Hong N. 3 hospital workers gave out masks. Weeks later, they all were dead. New York Times. Published online May 2020. https://www.nytimes.com/2020/05/04/nyregion/coronavirus-ny-hospital-workers.html Accessed 9 May 2020

